# Artificial Intelligence for Significant Mitral Regurgitation Screening and Diagnosis: A Systematic Review and Meta-analysis

**DOI:** 10.1101/2025.11.16.25340343

**Authors:** Udochukwu Godswill Anosike, Luena Seferasi, Marian Abedua Harrison, Ramiro Julian Nin Albonico, Sonia Ijeoma Etumudon, Leonardo Antunes Mesquita

**Affiliations:** Faculty of Medicine, Nnamdi Azikiwe University College of Health Sciences, Nigeria; Faculty of Medicine, University of Medicine Tirana, Tirana, Albania; Sakumono Specialist Hospital, Accra, Ghana; Centro Cardiológico Americano, Sanatorio Americano, Montevideo, Uruguay; Saratov State Medical University, Russia; Department of Electrophysiology, Hospital Madre Teresa, Belo Horizonte, Brazil

**Author notes:** **Disclosures:** All authors report no relationships that could be construed as a conflict of interest. All authors take responsibility for all aspects of the reliability and freedom from bias of the data presented and their discussed interpretation.

**Keywords:** Mitral regurgitation, Electrocardiogram, Echocardiogram, Artificial intelligence, Deep learning, Machine learning

## Abstract

**Objectives:** To evaluate performance of artificial intelligence (AI) models using electrocardiogram (ECG) and echocardiogram (ECHO) for predicting significant mitral regurgitation (MR).

**Materials and methods:** We performed a systematic review and meta-analysis of studies assessing AI models based on ECG or ECHO for detection of significant MR. Search was conducted in PubMed, Scopus, and Cochrane. Endpoints included: sensitivity, specificity of the models. Area under the summary receiver-operating characteristic curve (AUC) was calculated using a bivariate random-effects model.

**Results:** Fifteen studies (n = 2,470,826) were included: seven using ECG (n = 2,467,390) and eight using ECHO (n = 3,436). For AI-ECG models validated on external datasets, pooled sensitivity was 87.7% (95% CI 80.4 to 92.5) and specificity was 54.0% (95% CI 40.3 to 67.1), with an AUC of 81%. For AI-ECHO, sensitivity was 89.7% (95% CI 78.2 to 95.5) and specificity was 92.8% (95% CI 81.8 to 97.4), with an AUC of 96%.

**Conclusion:** AI models applied to ECG and ECHO demonstrate strong performance for detecting significant MR and may support clinicians’ diagnosis of MR. Clinical implementation, however, requires further validation and external testing across diverse populations.

## Introduction

Mitral regurgitation (MR) is one of the most prevalent valvular heart diseases globally - affecting up to 2-3% of general population, and increases with age, reaching approximately 10% in individuals over 75 years old.^1,2^ Early identification of MR by imaging is critical to prevent irreversible progression to heart failure, reducing morbidity and mortality, and enabling timely therapeutic intervention. The European Society of Cardiology (ESC) and the European Association for Cardio-Thoracic Surgery (EACTS) guideline for management of valvular heart disease recommend timely intervention for symptomatic severe MR due to association with worse outcomes.^3^

Conventional imaging (electrocardiogram) is not sensitive or specific for MR and has no role in MR screening due to poor sensitivity. Increasing efforts have been made by medical ultrasound experts, mathematicians, and computer scientists to promote the integration of ultrasound, medicine, and AI for mitral valve analysis, thereby improving the accuracy of ultrasonic diagnosis, reducing the misdiagnosis rate, shortening the reporting time, and meeting growing clinical needs.^4^ Artificial intelligence (AI), a technological driving force at present, has emerged as a promising strategy for scalable and accessible MR screening and diagnosis using electrocardiogram (ECG) and echocardiogram (ECHO).

AI-ECG can screen for moderate-to-severe valvular disease (including MR) with good discrimination, and modest positive predictive value in low-prevalence settings - so it is best used to prioritize ECHO.^5^ AI models are utilizing conventional 12-lead ECGs to detect early signs of significant MR - QRS complex alterations, abnormal P-wave and T-wave morphology, improving identification of undetected moderate-to-severe MR, predicting progression to atrial fibrillation or left ventricular dysfunction and accelerating patient referral for confirmatory echocardiography.^6,7,8^

AI-ECHO has the potential to automate view selection and quantification, decrease operator and inter-observer variability in image acquisition, measurements, and interpretation.^9^ For assessing mitral regurgitation (MR) severity, it has demonstrated feasibility, speed, and high diagnostic accuracy, with predictive capability for 1-year mortality.^10^ Clinical integration may enhance patient outcomes by streamlining referrals to specialized care and improving access to evidence-based treatments, while also optimizing workflow and diagnostic efficiency in echocardiography labs.

Therefore, we conducted a systematic review and meta-analysis to evaluate the diagnostic accuracy of AI-based models for non-invasive detection of moderate-to-severe or severe MR through ECG and ECHO, synthesize evidence across various modalities, algorithms, and identify key sources of heterogeneity, and validation quality. Findings from this study may help to translate AI to bedside and to enhance clinician screening and diagnosis of MR.

## Methods

### Search Strategy

This systematic review and meta-analysis were performed and reported in accordance with the Preferred Reporting Items for Systematic Reviews and Meta-Analyses of Diagnostic Test Accuracy (PRISMA-DTA) guidelines.^11^ The protocol was registered in the International Prospective Register of Systematic Reviews (PROSPERO) under CRD420251090360. PubMed, SCOPUS, and Cochrane Library were systematically searched from inception to June 2025. Boolean operators (AND, OR) and the following terms were used in the search strategy: “mitral regurgitation”, “mitral valve”, “artificial intelligence”, “AI”, “machine learning”, “deep learning”, “electrocardiogram”, “ECG”, “echocardiogram”, “ECHO”. References from all included studies were also manually searched for any additional eligible studies (backward snowballing).

### Inclusion and exclusion criteria

Studies were included if they met all the following criteria: (1) evaluated the use of AI models based on ECG or ECHO for detection of significant MR; (2) in pediatric and adult patients; and (3) reported any outcomes of interest.

We excluded studies if they (1) involved overlapping patient populations; (2) were case reports, reviews, letters, or abstracts. Two separate reviewers (U.G.A. and L.S.) conducted screening of titles and abstracts, and selected studies underwent full-text assessment based on inclusion and exclusion criteria. Discrepancies were resolved by a third reviewer (L.A.M.) in consensus. Significant MR represents patients with moderate or severe MR.

### Data extraction

Two reviewers (U.G.A. and L.S.) independently extracted data from the selected studies into a standardized form, and discrepancies were resolved by consensus with a third reviewer (L.A.M.). Following study characteristics were collected: (1) authors; (2) year of publication; (3) study design; (4) country, (5) imaging technique, (6) AI model, (7) Diagnostic performance of AI-ECG and AI-ECHO, including the number of true positives (TP), false positives (FP), false negatives (FN), and true negatives (TN), sensitivity and specificity. We extracted the following baseline patient data: (1) mean/median age; (2) percentage of male/female; (3) mean/median body mass index (BMI); (4) mean/median left ventricular ejection fraction (LVEF); (5) mean/median left atrial diameter (LAd); (6) percentage of patients with comorbidities (hypertension, diabetes, coronary artery disease, heart failure, atrial fibrillation).

### Quality assessment

To assess bias, each study included in the analysis was independently evaluated by two reviewers (L.A.M. and U.G.A.) using Quality Assessment of Diagnostic Accuracy Studies (QUADAS-2) tool *(Supplementary Figure 1*).^12^ This framework consists of four key domains: patient selection (D1), index test (D2), reference standard (D3), and flow and timing (D4). Each domain is analyzed for potential sources of bias and rated as low, high or unclear risk. Studies are categorized into one of two groups: “low risk of bias” if they received a low risk rating across all domains, or “at risk of bias” if any domain was rated as high or unclear risk. Discrepancies between the two reviewers were resolved by consulting a third reviewer (L.S.)

### Statistical analysis

The pooled diagnostic sensitivity and specificity with corresponding 95% confidence intervals (CIs) were obtained using a bivariate random-effects model. Estimates of sensitivity and specificity were summarized in a summary receiver-operating characteristic curve (SROC) and forest plots to provide a graphical overview of the outcomes and potential sources of heterogeneity. Area under the curve (AUC) for the SROC was estimated using parametric bootstrapping.^13^ Heterogeneity of studies was qualitatively assessed through visual examination of forest plots and quantitatively evaluated using the Zhou and Dendukuri bivariate Bayesian approach for diagnostic meta-analysis (I^2^ > 50% was considered significant heterogeneity).^14^ Correlation between logit sensitivity and 1-specificity was calculated to assess the potential for a threshold effect, and a coefficient (ρ) ≥ 0.6 was interpreted as significant.^15^ Subgroup analysis was performed based on pre-specified covariate (AI model). Including year as a covariate allows evaluation of temporal trends in diagnostic performance, reflecting advances in AI methods and datasets over time. Sensitivity analyses explored sources of heterogeneity, and Deeks’ test for publication bias was planned if ≥10 studies were included. Analyses were conducted in R (version 4.5.1).

## Results

### Study selection and characteristics

The initial search returned 930 studies, of which 94 were duplicates. We screened titles and abstracts of the remaining studies (n = 837). Of these, 30 studies were full text assessed for eligibility, and we excluded 15 studies. We included 15 studies, comprising 340,893 patients with significant MR and 2,126,497 with no MR or MR but mild [Fig. 1].

**Fig 1.**
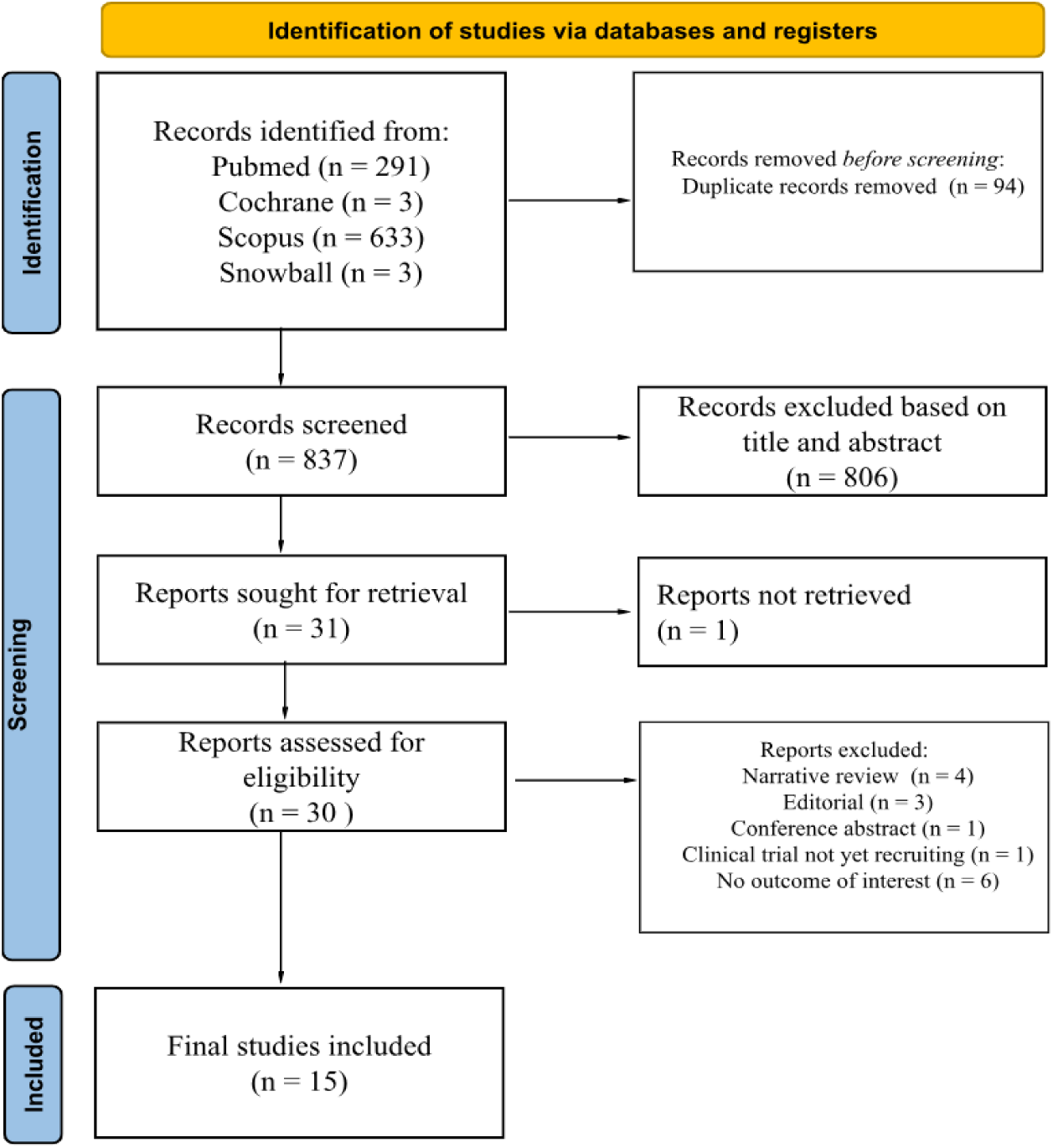
PRISMA FLOW DIAGRAM OF STUDY SCREENING AND SELECTION

All studies included were retrospective cohort. The median age of patients ranged from 64 to 71 years, 57% were male, and median LVEF ranged from 45% to 63%. A total of seven studies employed AI to predict MR using ECG^5,7,8,16,17–19^, while eight studies used ECHO^20–27^. Baseline characteristics were similar across significant MR and control group. Refer to Table 1 for baseline characteristics of the included studies and *supplementary table 1* for the key findings of each study.

**TABLE 1.** BASELINE CHARACTERISTICS OF STUDY.

|  | Country | Male (%) | Age (years) | LAd (mm) | DM (%) | HTN (%) | CAD (%) | HF (%) | AF (%) | LVEF (%) |
| --- | --- | --- | --- | --- | --- | --- | --- | --- | --- | --- |
| Vaid 2023 <sup>16</sup> | USA | 49.5 | 71.4 (71.3–71.5) | NA | NA | NA | NA | NA | NA | NA |
| Ulloa-Cerna 2022 <sup>17</sup> | USA | 50.1 | 64 (52-76) | NA | 23.10 | 46.40 | 23.10 | 17.20 | 8.5 | NA |
| Kwon 2020 <sup>8</sup> | South Korea | 40.4 | 67.74± 13.85 | 49.56±10.26 | NA | NA | NA | NA | 41.6 | 45.57± 11.95 |
| Lin 2024 <sup>5</sup> | Taiwan | 51.8 | 65.3±2.11 | 38.8±0.55 | 30 | 51.4 | 32.6 | 15.4 | 8.1 | 63.75±2.4 |
| Dhingra 2025 <sup>6</sup> | USA | 50.2 | 66.9 ± 15.5 | NA | NA | NA | NA | NA | NA | NA |
| Sakuma 2025 <sup>19</sup> | Japan | 62 | 66.1±14.9 | 46.4±9.9 | 21.10 | 63.90 | 3 | 30.40 | 13 | 56.1±18.4 |
| Shiraga 2023 <sup>18</sup> | USA | 55 | 67.0 ± 15.7 | NA | 20.70 | 65.50 | NA | NA | 17 | 58.7±10.4 |
| Vrudhala 2024 <sup>23</sup> | USA | 57 | NA | 36.88±0.55 | NA | 61.5 | 42.9 | NA | 32.6 | 56.7±11.1 |
| Yang 2022 <sup>24</sup> | China | 66.5 | 68.3 (59-79.3) | 40.3 (35-43.7) | 19.3 | 55.4 | 47.3 | 3.7 | NA | 54 (38-59) |
| Brown 2023 <sup>22</sup> | USA | NA | NA | NA | NA | NA | NA | NA | NA | NA |
| Moghaddasi 2016 <sup>21</sup> | Tehran | NA | NA | NA | NA | NA | NA | NA | NA | NA |
| Edwards 2022 <sup>20</sup> | Malawi | NA | NA | NA | NA | NA | NA | NA | NA | NA |
| Zhong 2024 <sup>27</sup> | China | 60 | 65.6 ± 11.2 | 48 (44, 52) | 16.9 | 41.5 | 56.8 | NA | 34.6 | 50 (39, 56) |
| Zhang 2021 <sup>25</sup> | China | 74.7 | 70.7 | 40.7±7.1 | 74.8 | 85.1 | 86.2 | NA | NA | 50.3 (22-31) |
| YangRP 2022 <sup>26</sup> | China | 68.7 | 69.5 ± 13.5 | 41 (36, 44) | 23 | 44.2 | 50.6 | 5 | NA | 53(37, 59) |

Continuous data are represented as mean/median and SD. Binary data are represented as percentage. LAd = left atrial diameter; DM = diabetes mellitus; HTN = hypertension; CAD = coronary artery disease; HF = heart failure; AF = atrial fibrillation; LVEF = left ventricular ejection fraction

### Quality appraisal

Study’s quality assessed by QUADAS-2 tool is illustrated in *supplementary Figure 1*. One study was classified as low risk, while the remaining 14 were unclear risk. Most studies were downgraded as unclear risk in “index test” domain due to unclear blinding between AI-based ECG interpretation and echocardiographic reference standard.

### Diagnostic accuracy of significant MR

#### Pooled analysis

The forest plots demonstrating the sensitivity and specificity of the included studies are shown in Fig 2&4. The correlation between logit-sensitivity and 1-specificity for AI-ECG and AI-ECHO were 0.93 and −1.00 respectively, indicating presence of a threshold effect. Due to inconsistent reporting of the diagnostic thresholds among the included studies, meta-regression analysis to explore the source of the threshold effect could not be performed. Consequently, pooled sensitivity and specificity were interpreted cautiously, and the hierarchical summary receiver operating characteristic (HSROC) model and area under the curve (AUC) which account for the trade-off between sensitivity and specificity, were used to summarize diagnostic performance.

**Fig 2.**
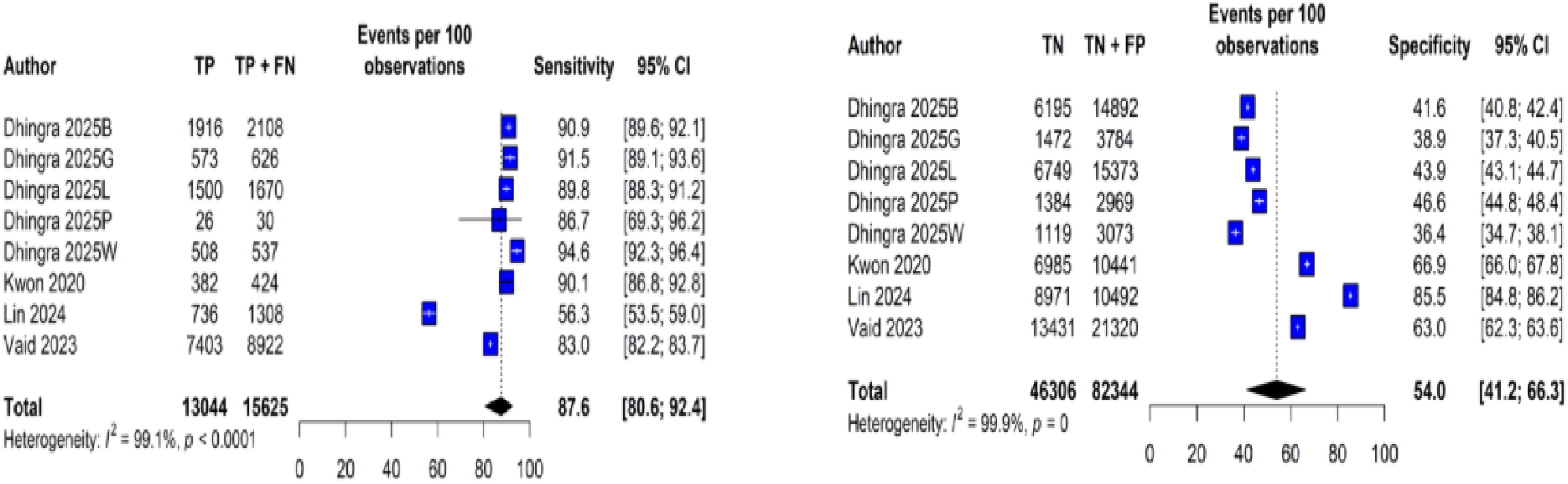
Forest plot of AI-ECG model external validation set (EVS) demonstrating sensitivity and specificity.

Using a bivariate approach, the pooled sensitivity and specificity of 87.7% of AI-ECG models were (95% CI 80.4 to 92.5) and 54% (95% CI 40.3 to 67.1), respectively (Fig 3). AUC for the pooled sROC curve was 81% (95% CI 76 to 85). There was substantial heterogeneity using the bivariate model (I^2^ = 93.5%), reflecting varied architectures and datasets..

**Fig 3.**
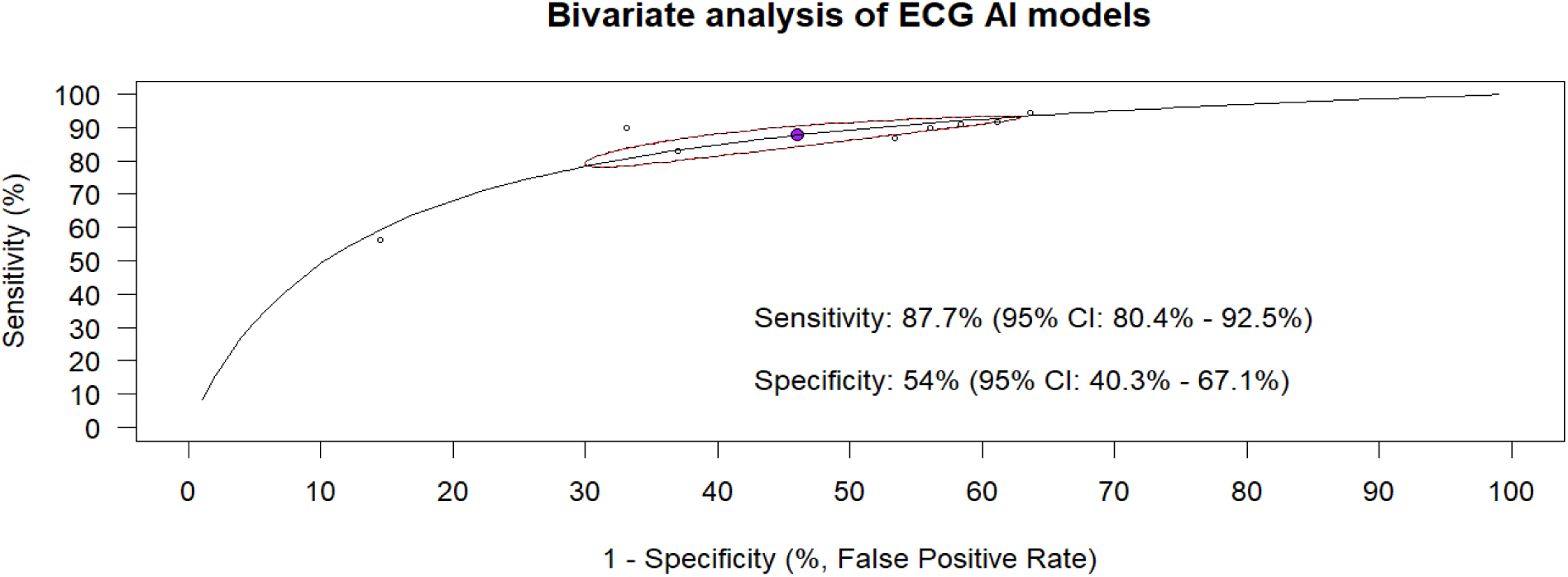
Summary Receiver-Operating Characteristic Curve (sROC) for ECG detecting significant mitral regurgitation. The filled dark circle represents the summary point of sensitivity and specificity. Each circle surrounding the sROC curve reflects one of the included studies with a pair of sensitivity and 1-specificity. The 95% confidence interval around the summary estimates represents a 95% probability the true average sensitivity and specificity of AI-ECG lies within that region. The area under the curve (AUC) was 0.81 (95% CI 0.76–0.85)

Using a bivariate approach, pooled sensitivity and specificity of AI-ECHO models were 89.7% (95% CI 78.2 to 95.5) and 92.8% (95% CI 81.8 to 97.4), respectively (Fig 5). AUC for the pooled sROC curve was 96% (95% CI 0.85 to 0.98). There was no heterogeneity using the bivariate model (I^2^ = 0%) - this likely reflects the narrow internal test design rather than absence of heterogeneity. Endpoints for AI-ECHO external validation set were not available in studies.

Dhingra et al. reported individual endpoints of sites where AI-ECG model was internally tested and externally validated (*Fig 2*). B, G, L, W, P represent external validation hospital sites. B - Bridgeport hospital; G - Greenwich hospital; L - Lawrence + Memorial hospital; W - Westerly hospital; P - Population-based cohort (ELSA-Brazil). Vrudhala et al. and Moghaddasi et al. reported individual endpoints of MR severity and machine learning models respectively (*Fig 4*). M - Moderate MR; S - Severe MR. MS - Support vector machine model; ML - Linear discriminant analysis model + convolutional neural network.

**Fig 4.**
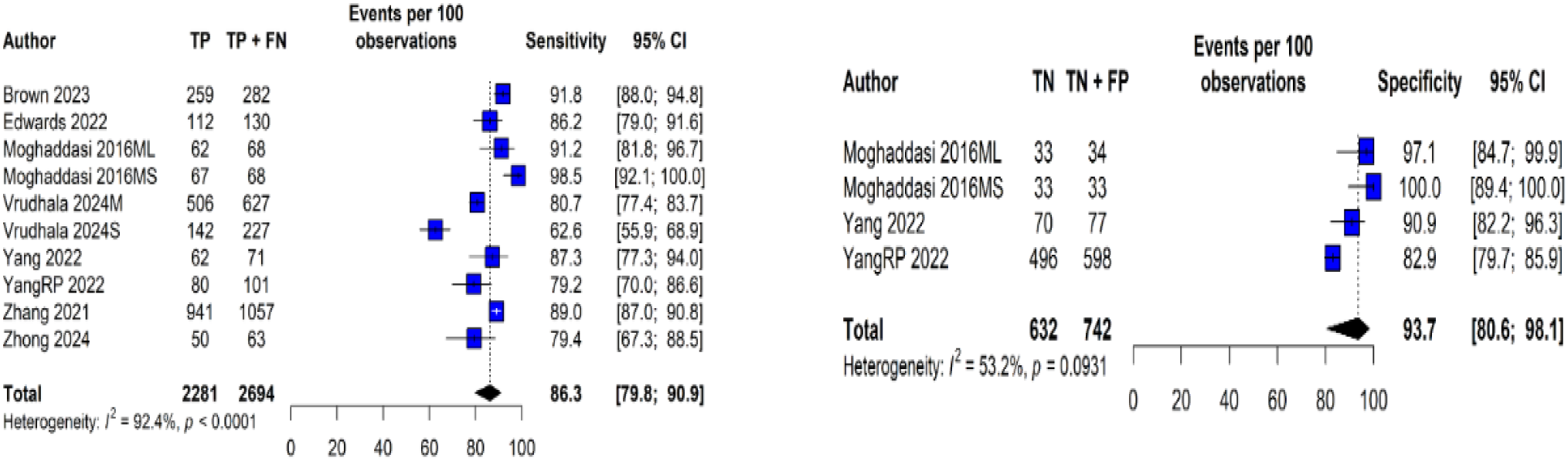
Forest plot of AI-ECHO models internal test set (ITS) demonstrating sensitivity and specificity.

**Fig 5.**
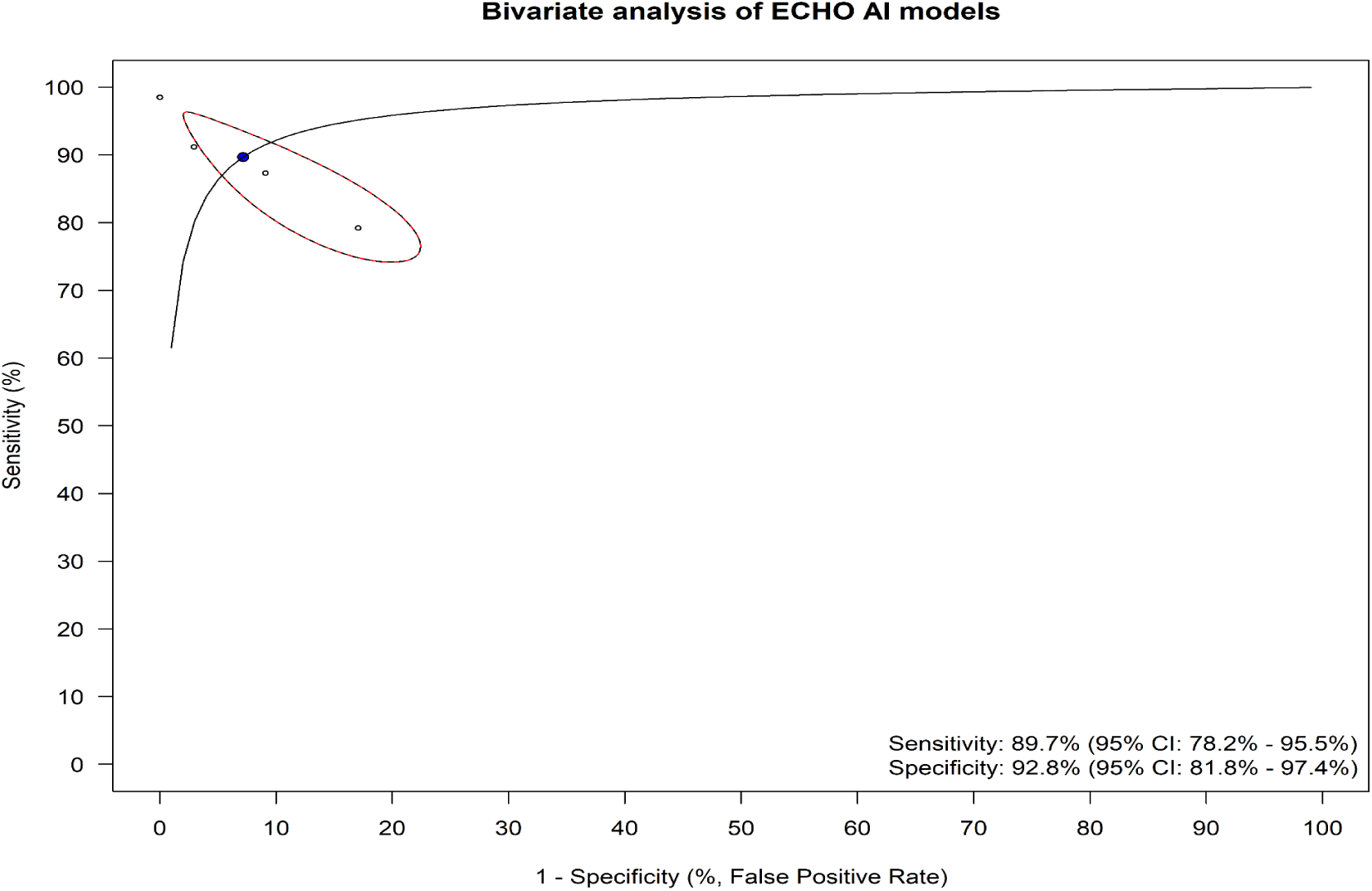
Summary Receiver-Operating Characteristic Curve (sROC) for ECHO detecting significant mitral regurgitation. The filled dark circle represents the summary point of sensitivity and specificity. Each circle surrounding the sROC curve reflects one of the included studies with a pair of sensitivity and 1-specificity. The 95% confidence interval around the summary estimates represents a 95% probability the true average sensitivity and specificity of AI-ECHO lies within that region. The area under the curve (AUC) was 0.96 (95% CI 0.85–0.98)

#### Subgroup analysis

Subgroup analysis compared the performance of AI models applied to ECHO (***Fig 6***). Machine learning (ML) showed better performance than deep learning for diagnosing significant MR - though only very few studies were involved.^17,20,21^ Deep learning was the major AI model used in ECG studies - hence the absence of subgroup analysis.

**Fig 6.**
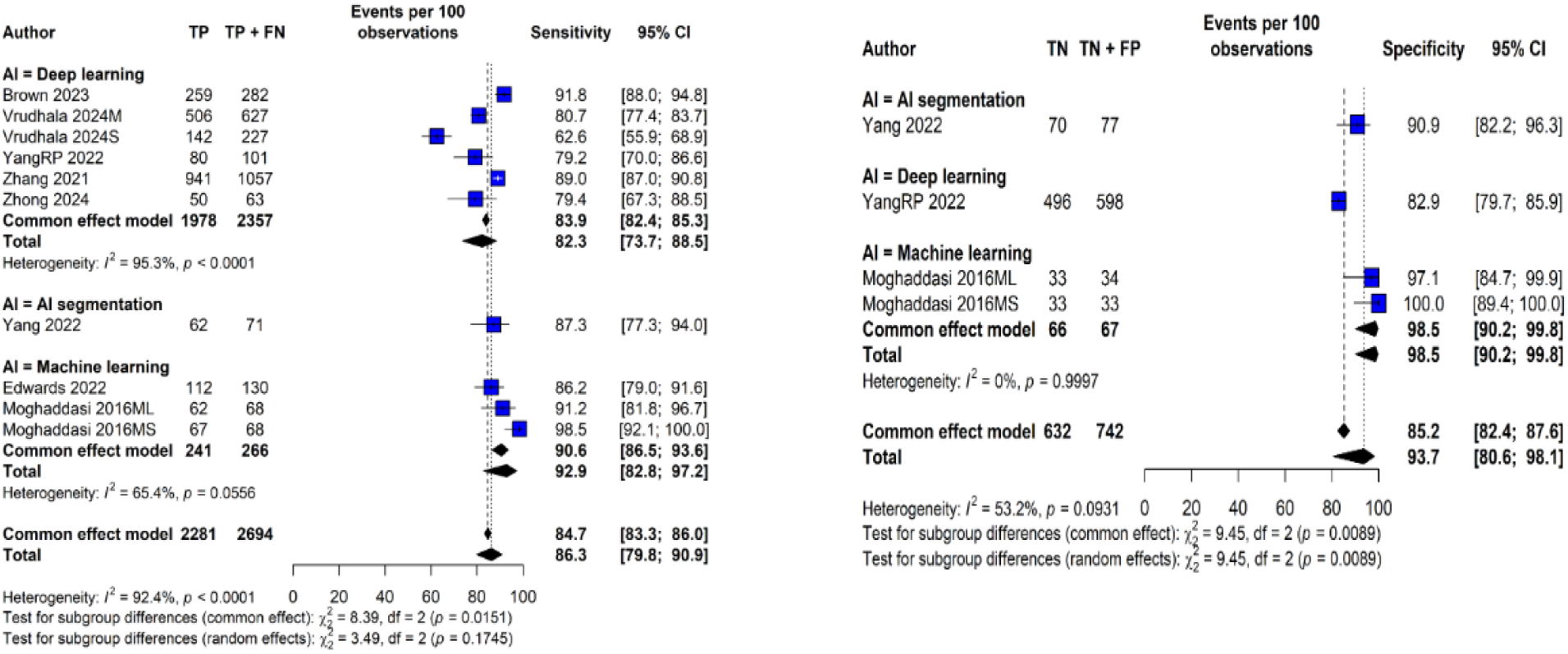
Subgroup analysis of AI-ECHO model demonstrating sensitivity and specificity.

#### Sensitivity analyses

The leave-one-out analysis *(supplementary Table 3)* showed consistent pooled estimates for both sensitivity and specificity, indicating the robustness of the findings. Notably, exclusion of Shiraga et al. (2023) led to the largest increase in pooled sensitivity, indicating that this study contributed a lower-than-average sensitivity. Exclusion of Dhingra et al. (2025) and Kwon et al. (2020) resulted in a modest increase in specificity. No individual study significantly altered the overall estimates, suggesting minimal influence from outliers and stable diagnostic performance of the AI-ECG models.

## Discussion

This systematic review and meta-analysis included 15 studies with a total of 2,470,826 patients. Of these, seven studies were based on ECG studies (n = 2,467,390) and eight were based on ECHO (n = 3,436). Our analysis demonstrated that AI models based on ECG showed high sensitivity (88%) but limited specificity (54%) for detecting significant MR. In contrast, models based on ECHO showed excellent specificity (93%) and a similarly high sensitivity (90%).

Sensitivity ≥87% suggest that AI-ECG models could be clinically useful as screening tools for significant MR. This could help triage patients for further echocardiographic evaluation, especially where access to imaging is limited. On the other hand, the low specificity of AI-ECG may lead to increased false positives, with unnecessary downstream testing if used in isolation, and serves as a barrier to clinical implementation as a screening tool. Furthermore, the introduction of a standardized flowchart (for concomitant assessment of symptoms or comorbidities that increase the pre-test probability of MR diagnosis) to support AI-ECG at the initial stage of screening should be considered as this may reduce the rate of false positives as well as unwarranted echocardiogram. As shown in *supplementary table 2*, convolutional neural network was the prominent architecture employed by studies to aid AI models process data from ECGs with accuracy and precision.

There were differences in the sensitivity and specificity among the included studies. Heterogeneity of estimates using the bivariate model was also significant. Upon further exploration, studies by Lin et al., and Vaid et al., contributed the most to the heterogeneity based on inspection of EVS forest plots (*Fig 2*) with relatively lower sensitivity and higher specificity than others.^16,18^ Variability in comorbidities, and clinical setting likely contributed to the observed heterogeneity. Notably, Lin et al.’s external cohort included older patients with more comorbidities and combined data from an academic medical center and a community hospital, introducing heterogeneity in population characteristics and clinical practice. Likewise, Vaid et al. reported significant demographic differences in external cohorts. These findings underscore the importance of contextual model validation and the need for external datasets that reflect real-world diversity to ensure robust and generalizable AI performance. The implementation of noninvasive and scalable AI-based ECG as a screening tool could improve early detection of mitral regurgitation and facilitate timely referral and management.

While cardiologist interpretation of echocardiography is essential, the low resolution and artifacts in images can hinder decision-making, making computer-aided diagnosis (CAD) particularly valuable.^21^ AI-ECHO automates quantification of MR severity, provides mechanistic classification and informs treatment - which AI-ECG cannot offer. Our study highlights the high sensitivity and specificity observed in AI-ECHO models signaling their clinically viability and ability to support clinicians as a tool for accurate diagnosis of significant MR. Several architectures (video-based convolutional neural network, self-supervised learning algorithm, support vector machine, fully convolutional neural network and mask region with convolutional neural network) were employed by studies to aid AI models process data from ECHOs with accuracy and precision (*Supplementary table 2*). According to our subgroup analyses (*Fig 6*), machine learning models outperformed deep learning models. This contrasts with a study assumption that deep learning approaches have shown superior performance compared to machine-learning (ML) approaches when applied to ECHO.^9^ This apparent superiority of ML over DL in ECHO-based studies should be interpreted cautiously, given the very small number of ML studies and potential differences in dataset size and quality.

Though focused cardiac ultrasound (FCU), a form of POCUS can provide reasonably accurate assessments of chamber size, ventricular function, pericardial effusion, and central venous volume, it is suboptimal for evaluating more complex cardiac pathology such as valvular disease and diastolic dysfunction.^28^ The accuracy of fully automated ML-enabled POCUS devices for quantification of LVEF were comparable to reference echocardiograms interpreted by expert cardiologists.^29^ Beyond quantification of LVEF, ML-enabled POCUS and DL-enabled POCUS can become useful diagnostic tools for accurate detection of significant MR. Therefore, the integration of artificial intelligence with POCUS can move cardiac ultrasound beyond the echo lab, improving access to efficient, high-quality care.

Based on the total sample, the database size of AI-based ECG studies was evidently larger than AI-based ECHO studies (*supplementary table 1*). The larger database size of AI-based ECG studies likely reflects practical, logistical, and technical factors - ECGs are cheaper, standardized, widely available, easy to digitize, and easier to share than ECHOs. The implication is that ECG AI models are more likely to reach real-world clinical deployment faster because large datasets support robust validation whereas ECHO AI models may lag behind, needing multicenter collaborations to achieve sufficient data volume. Also, majority of AI-ECG studies utilized complex deep learning model architecture such as convolutional neural networks while AI-ECHO studies had variations in the model architecture used (*supplementary table 2*). Despite the variations in size of datasets and model architecture, included studies reported strong performance across diverse patient demographics including those at risk of developing moderate-to-severe or severe MR (pediatric patients with rheumatic heart disease and adult patients with chronic atrial fibrillation).

Quality assessment using the QUADAS-2 tool (*supplementary figure 1*) revealed that most included studies were at unclear risk of bias, primarily due to insufficient reporting of blinding between the AI model output and the reference standard. Only one study was rated as low risk across all domains. The most frequent concern was in the index test domain, where lack of clarity regarding interpretation independence was common. These findings highlight the methodological limitations in the current literature and reinforce the need for standardized reporting and rigorous design, including prospective validation and clear blinding protocols, in future AI diagnostic studies.

Our study has several limitations. We included a few studies, only seven AI-ECG and eight AI-ECHO studies met the eligibility criteria, which may limit the overall generalizability of our findings. Existing studies vary widely in AI architecture used and range of performance (*Supplementary table 2*). Meta-regression based on age group, disease etiology (degenerative/functional vs rheumatic), and MR grading could not be performed due to limited data. Also, the inability to conduct meta-regression due to insufficient threshold data restricted the evaluation of threshold-related heterogeneity. This limitation may affect the precision of pooled estimates and underscores the importance of consistent threshold reporting in diagnostic research. Notably, none of the AI-ECHO studies reported external validation datasets, restricting the ability to assess model performance across independent populations and real-world settings. Among the AI-ECG studies, only four included external validation cohorts, which limits the strength of conclusions. Fewer studies were available for the estimation of specificity compared to sensitivity in analyses of AI-ECHO models. This imbalance may introduce bias in the pooled specificity estimate and limit its precision. Additionally, studies that reported specificity may differ methodologically or clinically from those that did not, potentially affecting the representativeness of the findings. As a result, conclusions drawn regarding the ability of AI-augmented ECHO to correctly identify patients without significant MR should be interpreted with caution.

## Conclusion

In this systematic review and meta-analysis, we found that AI-based tools leveraging ECG and echocardiographic data show promise in predicting significant mitral regurgitation (MR). AI-ECG models offer high sensitivity but limited specificity, suggesting potential as a screening tool, while AI-ECHO demonstrates both high sensitivity and specificity, supporting its use in diagnostic workflows. Despite encouraging results, limited generalizability and low specificity of AI-ECG pose challenges for clinical implementation. Standardized datasets and external validation are needed to advance clinical adoption and integration into guidelines for diagnosis of MR.

## Data Availability

All data produced in the present study are available upon reasonable request to the authors

## ABBREVIATIONS

AI: Artificial Intelligence
AUC: Area under curve
CMR: Cardiac Magnetic Resonance
DL: Deep learning
ECG: Electrocardiogram
ECHO: Echocardiography
EVS: External validation set
FCU: Focused cardiac ultrasound
ITS: Internal test set
ML: Machine learning
MR: Mitral regurgitation
POCUS: Point-of-care-ultrasound
PRISMA: Preferred Reporting Items for Systematic Review and Meta-Analyses
PROSPERO: International Prospective Register of Systematic Reviews
QUADAS: Quality Assessment of Diagnostic Accuracy Studies
sROC: Summary receiver-operating characteristic curve

