## Supplementary Material for "Artificial Intelligence for Significant Mitral Regurgitation Screening and Diagnosis: A Systematic Review and Meta-analysis"

**Supplementary Table 1.** Number of patients with/without MR, prevalence, definitions, methods of assessment and key findings of included studies.

**Supplementary Table 2.** Descriptions of AI models, database size, method/architecture used, range of performance in included studies for MR.

**Supplementary Table 3.** Leave-One-Out analyses of Sensitivity and Specificity Results of AI-ECG studies.

**Supplementary Figure 1.** QUADAS-2 risk of bias assessment of included ECG and ECHO studies

**Supplementary Table 1.** Number of patients with/without MR, prevalence, definitions, methods of assessment and key findings of included studies

|  | **Total sample** (with/without MR) | **MR prevalence (%)**  internal testing/external validation | **MR definition** | **MR assessment methods** | **Primary aim/Key findings** |
| --- | --- | --- | --- | --- | --- |
| **Vaid** 2023 ^16^ | 607,429 | 7.11/7.32 | Moderate-to-severe or severe | employed Natural Language Processing (NLP) to extract relevant data from echocardiogram reports. ECG data were exported from  the GE MUSE ECG system as structured. | **Aim**: explored the application of deep learning algorithms to ECGs from a diverse population for identifying left heart valvular dysfunction, specifically Aortic Stenosis and Mitral Regurgitation  **Key finding**: deep learning models demonstrated strong performance across diverse patient demographics and clinical scenarios |
| **Ulloa-Cerna** 2022 ^17^ | 1,748,940 | 4.5/NR | Defined as Moderate or severe based on echocardiographic reports | extracted echocardiographic measurements and diagnoses from Xcelera reports and ECG structured findings,  measurements, and 12-lead traces from MUSE | **Aim**: to improve diagnostic screening in patients at increased risk of undiagnosed structural heart disease using ECG-based machine learning model  **Key finding:** ECG-based machine learning model effectively identifies patients at high risk for undiagnosed structural heart disease, enhancing screening recommendations for echocardiography |
| **Kwon** 2020 ^8^ | 14,039 | 25.96/3.90 | Significant MR (moderate  to severe), defined as effective regurgitant orifice area ≥ 0.2 cm2, regurgitation  volume ≥ 30 ml, regurgitation fraction ≥ 30%, and MR grade II–IV confirmed by echocardiography | sensitivity map was employed to visualize which parts of the ECG contributed most to the AI's decision-making, revealing a focus on the P-wave and T-wave for MR patients | **Aim**: to leverage AI to enhance the detection of MR through standard ECG  **Key finding:** AI algorithm demonstrated strong performance in detecting MR, and effectively identified high-risk patients who later developed MR |
| **Lin** 2024 ^5^ | 19,886 | 10.8/11.1 | moderate mitral regurgitation was defined as a central jet MR of 20–39%, a vena contracta of 0.3–0.69 cm, a regurgitant volume of 30–59 mL, a regurgitant fraction of 30–49%, or an ERO 0.2–0.39 cm2, and the definition of severe mitral regurgitation was a central jet MR >40%, a vena contracta ≥0.7 cm, a regurgitant volume ≥60 mL, a regurgitant fraction ≥50%, or an ERO ≥0.40 cm2 | MR was assessed using comprehensive 2D echocardiograms recorded on a Philips imaging system | **Aim**: to explore the effectiveness of artificial intelligence-enabled electrocardiograms (AI-ECG) in screening for multiple valvular heart diseases (VHDs)  **Key finding:** AI-ECG models demonstrated promising performance in identifying various valvular diseases, with notable sensitivity and specificity |
| **Dhingra** 2025 ^6^ | 55,326 | 9.3/52.3 | Did not clearly define MR – however, included moderate-to-severe MR as part of the composite structural heart disease (SHD) outcome. | MR was assessed using transthoracic echocardiograms (TTEs) interpreted by board-certified cardiologists according to established echocardiographic guidelines. | **Aim**: presents an innovative approach using deep learning algorithms to screen for structural heart diseases (SHDs) through electrocardiographic images.  **Key finding:** The model's performance was consistent across demographic subgroups and various ECG formats. |
| **Sakuma** 2025 ^19^ | 19,170 | 2.9/NR | defined as moderate or severe MR | MR was assessed using echocardiography | **Aim**: to explores the effectiveness of AI-algorithm using ECG dataset in diagnosing and predicting mitral regurgitation (MR) in patients with chronic atrial fibrillation (AF)  **Key finding:** the INN model outperformed the initial CNN model in detecting MR |
| **Shiraga** 2023 ^18^ | 212 | 18.8/NR | defined based on echocardiographic guidelines. The specific criteria for grading MR severity were not detailed | MR was assessed using a combination of auscultation and ECG data | **Aim**: to evaluate the effectiveness of combining auscultation and electrocardiography data with machine learning to improve the diagnosis of valvular pathologies and ventricular dysfunction  **Key finding:** combining auscultation and electrocardiography (ECG) data using multimodal artificial intelligence (AI) significantly improves the diagnostic efficiency for detecting severe aortic stenosis (AS), severe mitral regurgitation (MR), and left ventricular dysfunction (LVD). |
| **Vrudhala** 2024 ^23^ | 4,128 | 41.4/14.4 | defined based on clinical echocardiogram reports following American Society of Echocardiography guidelines | The pipeline automatically identified apical 4-chamber (A4C) view videos with color Doppler across the mitral valve from a large dataset of echocardiograms | **Aim**: to detect clinically significant MR from transthoracic echocardiograms using a deep learning pipeline  **Key finding:** AI model effectively distinguished between different severities of MR, showcasing strong performance metrics |
| **Yang** 2022 ^24^ | 148 | 48/NR | Moderate and severe MR were considered as positive disease status, while non-moderate or severe MR was considered as negative. | MR severity was assessed using a combination of clinical echocardiographic reports and quantitative measurements based on the 2017 ASE Recommendations for Noninvasive Evaluation of Native Valvular Regurgitation | **Aim**: explores the effectiveness of an AI segmentation model in assessing the severity of mitral regurgitation (MR) using Color Doppler echocardiography.  **Key finding:** The integration of AI significantly enhanced the diagnostic accuracy of physicians interpreting echocardiographic studies |
| **Brown** 2023 ^22^ | 511 | NR | Pathologic MR was defined based on the 2012 World Heart Federation (WHF) criteria for echocardiographic diagnosis of RHD | MR was assessed by interpretation of spatiotemporal information from echocardiographic images using deep learning models, and assessing  proximity of maximum MR jet to the mitral valve and its blue  color intensity using machine learning | **Aim**: employed machine learning and deep learning techniques to analyze MR jets and detect RHD in children, using convolutional neural networks (CNNs) and attention mechanisms  **Key finding:** machine learning and deep learning approaches can effectively identify RHD, paving the way for enhanced screening strategies globally. |
| **Moghaddasi** 2016 ^21^ | 203 | NR | severity of mitral regurgitation was graded qualitatively as mild, moderate and severe according to the ACC/AHA guidelines for valvular heart  disease | employed two novel features: Extensive Local Binary Pattern (ELBP) and Extensive Volume Local Binary Pattern (EVLBP) to analyze echocardiography images. severity of MR was evaluated based on textural features extracted from echocardiography images | **Aim**: to introduce a method for the automatic assessment of mitral regurgitation (MR) severity using 2D echocardiography videos, image processing techniques, and machine learning  **Key finding:** automatic assessment of MR severity using 2D echocardiography videos is a promising approach that combines advanced image processing and machine learning techniques |
| **Edwards** 2022 ^20^ | 182 | NR | MR was defined as the presence of any regurgitation detected in systolic frames of the parasternal long-axis view with color Doppler (PLAX-C) | (MR) was assessed using a machine learning pipeline consisting of two convolutional neural networks (CNNs). The MR detection model relied heavily on the color Doppler signal and focused on systolic frames to ensure consistent data features | **Aim**: explores the development of a machine learning model to automate the detection of mitral regurgitation (MR) in pediatric echocardiograms  **Key finding:** Demonstrated feasibility for automated MR detection in rheumatic heart disease (RHD) screening |
| **Zhong** 2024 ^27^ | 121 | NR | MR was defined and graded based on the 2017 American Society of Echocardiography (ASE) guidelines, using quantitative parameters derived from the proximal isovelocity surface area (PISA) method | MR was assessed using transthoracic echocardiography (TTE) with the proximal isovelocity surface area (PISA) method | **Aim:** to automate the grading of MR severity using echocardiographic images  **Key finding:** The model demonstrated high accuracy in identifying MR severity |
| **Zhang** 2021 ^25^ | 1,427 | NR | MR was defined and classified based on the 2017 American Society of Echocardiography (ASE) guidelines. | The LabelMe software was utilized to mark regions of interest in the echocardiography images for subsequent analysis | **Aim:** to evaluate the feasibility and accuracy of using the Mask R-CNN algorithm for the automatic assessment of mitral regurgitation (MR) severity through color Doppler echocardiography images  **Key finding:** The model demonstrated high accuracy in classifying MR severity providing a reliable and efficient alternative to traditional echocardiographic methods |
| **YangRP** 2022 ^26^ | 699 | NR | MR was defined based on the appearance of color Doppler signals in the region of the left atrium during systole in apical 4-chamber–mitral valve–color Doppler imaging (A4C-MV-CDI) views | MR was assessed using color Doppler echocardiographic videos, and the severity of MR was quantified by calculating the ratio of the MR jet area to the left atrial (LA) area | **Aim: to** present a deep learning framework designed to automatically analyze echocardiographic videos for diagnosing valvular heart diseases (VHDs).  **Key finding:** The deep learning algorithm demonstrated high accuracy in classifying various valvular heart diseases |

**Supplementary Table 2.** Descriptions of AI models, database size, method/architecture used, range of performance in included studies for MR.

|  | **AI model** | **Model architecture** | **Software/hardware** | **Range of performance** |
| --- | --- | --- | --- | --- |
| **Vaid** 2023 ^16^ | Deep learning | utilized a combination neural network consisting of a Multi-Layer Perceptron (MLP) joined to an Efficientnet Convolution Neural Network (CNN) utilizing a joint fusion strategy. | Models were  trained on an Azure Cloud virtual machine on 4x NVIDIA v100  GPUs with 16GB VRAM each. | Model performance was strong in the internal testing dataset with an AUROC of 0.88 (95% CI: 0.88–0.89). This performance lowered to 0.81 (95% CI: 0.80–0.82) in the external validation  Dataset. In either of internal testing or external validation, AUROC was seen to be constant across groups based on race, age, and sex |
| **Ulloa-Cerna** 2022 ^17^ | Machine learning | developed a deep convolutional  neural network (CNN) consisting of 6 1-dimensional CNN-Batch Normalization-ReLU layer blocks, followed by a multilayer perceptron and a final logistic output layer | Software and Hardware were not specified. However, XGBoost was used as part of the classification pipeline for the final model | Area Under Receiver Operating Curve (AUROC): 0.911. Positive Predictive Value (PPV): 15.2% (at 90% sensitivity). Negative Predictive Value (NPV): 99.4% (at 90% sensitivity). Specificity: 76.4% (at 90% sensitivity) |
| **Kwon** 2020 ^8^ | Not specified | AI algorithm was developed using a convolutional neural network (CNN) with 2-dimensional convolution, max pooling, flatten, batch normalization, and dropout layers | TensorFlow (Google LLC, MountainView, CA USA) open-source software library was used. Hardware not specified | during the internal validation, the AUROC of the AI algorithm was 0.816 (95% confidence interval [CI], 0.811–0.820). During the external validation, AUROC of the algorithm were 0.877 (95% CI: 0.870–0.883) |
| **Lin** 2024 ^5^ | Deep learning | used a convolutional neural network (CNN) architecture | Not specified | high negative predictive values and AUCs above 0.8, making it a reliable tool for detecting moderate-to-severe mitral regurgitation |
| **Dhingra** 2025 ^6^ | Deep learning | used CNN models built upon the EfficientNet-B3 architecture, which has 384 layers and over 10million trainable parameters. | Adam optimizer was used for training the CNN models. XGBoost was used for the ensemble learning strategy to combine the outputs of the CNN models. Hardware not specified | model performance for detecting moderate or severe mitral regurgitation (MR) is as follows: AUROC (Area Under the Receiver-Operating Characteristic Curve): 0.792 (95% CI: 0.776-0.807). performance was consistent across external validation cohorts and real-world ECG image modalities |
| **Sakuma** 2025 ^19^ | Not specified | used a convolutional neural network (CNN) and integrated  neural network (INN) | constructed a CNN using the Keras Framework with a TensorFlow (Google; Mountain View, CA,  USA) backend and Python. model was trained on a computer with 128-GB RAM and single Quadro P-2200 (NVIDIA) graphics processing unit. | For CNN - Area Under the Curve (AUC) for the Receiver Operating Characteristic (ROC) curve: 0.836. Sensitivity: 0.725. Positive Predictive Rate (PPR): 0.164.  For INN - AUC for the ROC curve: 0.848. Sensitivity: 0.726. PPR: 0.157 |
| **Shiraga** 2023 ^18^ | Not specified | used a 10-layer CNN | stacking process using  Random Forest and XGBoost was performed using scikit-learn 0.23.2 and xgboost 1.6.2. hardware not specified | Precordial-lead ECG (PCECG): AUC = 0.801 |
| **Vrudhala** 2024 ^23^ | Deep learning | Video-based convolutional  neural networks (R2+1D) were used for view classification and  MR severity assessment. | using an ADAM optimizer, an initial learning rate of  1e-2, and a batch size of 24 on 2 NVIDIA RTX 3090 graphic processing units. | In the internal test cohort, MR moderate or greater in severity was detected with an AUC of 0.916 (0.899–0.932) and severe MR was detected with an AUC of 0.934 (0.913–  0.953). In the external test cohort, the model detected MR moderate or greater in severity with an AUC of 0.951  (0.924–0.973) and severe MR with an AUC of 0.969 (0.946–0.987). |
| **Yang** 2022 ^24^ | Not specified | Color doppler self-supervised learning algorithm based on a 2D ResNet-34 architecture | The model used Philips echocardiography machines (iE-elite and 7C) equipped with transducers S5-1 and X5-1. For manual segmentation, the software LabelMe was utilized | When physicians used the AI model's output to assist in grading MR severity: Sensitivity increased from 77.0% (without AI) to 86.7%. Specificity remained largely unchanged, at 91.5% (without AI) vs. 90.5% (with AI) |
| **Brown** 2023 ^22^ | Deep learning  Machine learning | Used convolutional neural network  Linear support vector machines | used Keras (version 2.6.0)  and TensorFlow (version 2.6.2) frameworks, and the  model training was performed on a desktop computer equipped with a GeForce GTX TITAN X GPU from NVIDIA (Santa Clara, CA), which has 12 GB of memory | The machine learning model based on MR jet analysis shows high accuracy in detecting RHD, with an AUC of 0.93 |
| **Moghaddasi** 2016 ^21^ | Machine learning | Support Vector Machine (SVM), Linear Discriminant Analysis (LDA) and Template Matching techniques are used as  classifiers to determine the severity of MR based on textural descriptors. | Not specified | Achieved 99.38% sensitivity and 99.63% specificity for the detection of the severity of MR |
| **Edwards** 2022 ^20^ | Machine learning | Convolutional neural network (CNN) models were developed, inspired by DenseNet27 and ResNet,28 with hyperparameter tuning | Python (version 3.5) with the Keras library (version 2.0.9) and TensorFlow back end (version 1.3.0). Nvidia GTX 1080TI graphics card for training the models | achieved testing accuracy  of 0.86 and an area under the receiver operating characteristic curve of 0.91. |
| **Zhong** 2024 ^27^ | Deep learning | Deeplabv3+ fully convolutional neural network (FCN) | Operating System: Linux Ubuntu 18.04.5 LTS (developed by Canonical Ltd., London, UK) Deep Learning Frameworks:  PaddlePaddle-GPU (version 2.4.2, developed by Baidu, Inc., Beijing, China). GPU**:** NVIDIA GeForce RTX 3090 (24GB, developed by NVIDIA Corporation, Santa Clara, CA, USA) | The model demonstrated high accuracy and reliability, with better performance for Grades I and IV compared to Grades II and III |
| **Zhang** 2021 ^25^ | Deep learning | mask regions with a convolutional neural network (Mask R-CNN) | LabelMe Software (Version 3.167): Used for image annotation. Google TensorFlow Framework:  Used to construct the network architecture of the Mask R-CNN algorithm. Hardware not specified | The model demonstrated high accuracy for both severity and grading classifications.  Accuracy:  Mild: 0.90, Moderate: 0.89, Severe: 0.91. Accuracy:  Grade I: 0.90, Grade II: 0.87, Grade III: 0.81, Grade IV: 0.91 |
| **YangRP** 2022 ^26^ | Deep learning | Not specified | Analyses were performed using algorithms written  in Python 3.6 software from the libraries of Numpy, Pandas, and Scikit-learn. The DL model was trained and tested on a server equipped with:  GPU: NVIDIA Tesla V100. CPU: Intel Xeon Gold 6248 | Retrospective Test Dataset: Area Under the Curve (AUC): 0.97 (95% CI: 0.95–0.99), Sensitivity: 0.94 (95% CI: 0.90–0.97), Specificity: 0.94 (95% CI: 0.90–0.97), Accuracy: 0.94 (95% CI: 0.91–0.97)  Prospective Test Dataset: AUC: 0.88 (95% CI: 0.86–0.90), Sensitivity: 0.79 (95% CI: 0.76–0.82), Specificity: 0.83 (95% CI: 0.81–0.86), Accuracy: 0.81 (95% CI: 0.79–0.83) |

**Supplementary Table 3.** Leave-One-Out analyses of Sensitivity and Specificity Results

| Left Out Study | Pooled Sensitivity (95% CI) | Pooled Specificity (95% CI) |
| --- | --- | --- |
| Kwon 2020 | 0.736 (0.554-0.917) | 0.745 (0.630-0.861) |
| Ulloa-cerna 2022 | 0.736 (0.554-0.917) | 0.706 (0.573-0.839) |
| Shiraga 2023 | 0.827 (0.723-0.931) | 0.690 (0.568-0.811) |
| Vaid 2023 | 0.728 (0.553-0.904) | 0.719 (0.585-0.854) |
| Lin 2024 | 0.784 (0.603-0.964) | 0.690 (0.568-0.812) |
| Dhingra 2025 | 0.738 (0.555-0.921) | 0.754 (0.652-0.855) |
| Sakuma 2025 | 0.765 (0.576-0.954) | 0.701 (0.570-0.832) |

**Supplementary Figure 1.** QUADAS-2 risk of bias assessment of included ECG and ECHO studies

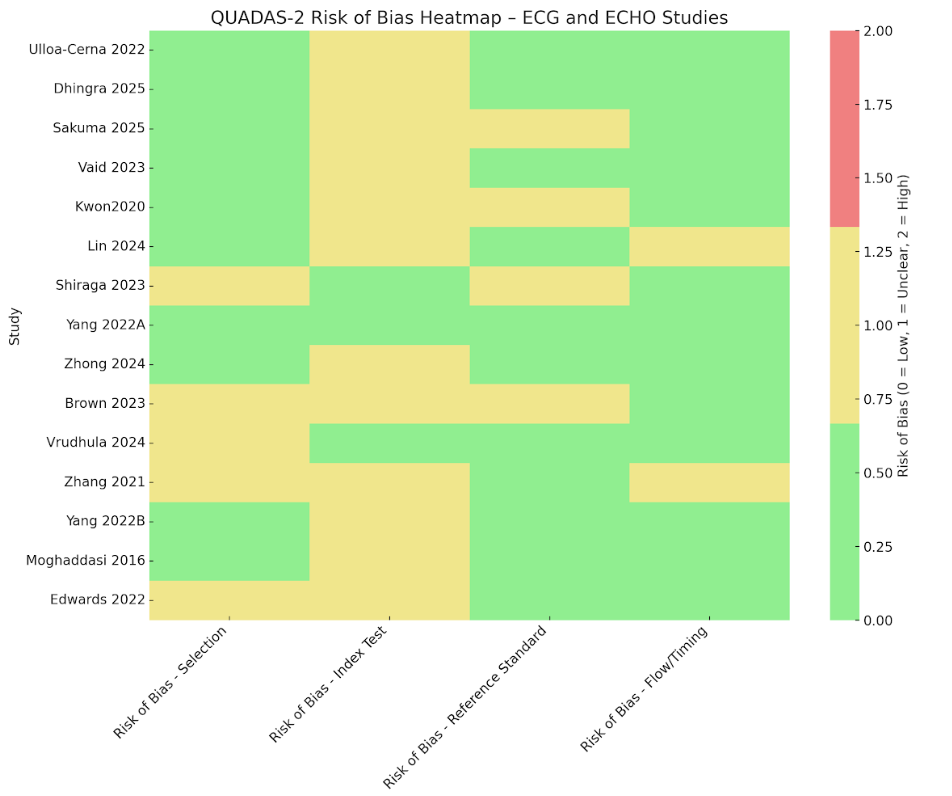
